# Evaluation of Device Quality and Satisfaction at Delivery during an Intensive Prosthesis Fitting Camp, and 3-Month Follow-Up

**DOI:** 10.1101/2025.06.04.25328969

**Authors:** Thearith Heang, Sisary Kheng, Maggie Donovan-Hall, Amos Channon, Alex Dickinson, Carson Harte

## Abstract

Prosthetic limbs enable mobility, independence, and social participation. Fitting camps provide mobility assistive devices to many individuals in intensive sessions, but concerns exist regarding device design, fabrication quality, and the required rehabilitation and aftercare.

This study evaluated a fitting camp in Cambodia where 525 people received prosthetic limbs. Using a multiple-methods study, we assessed device quality and client satisfaction at provision and at a 3-month follow-up structured interview.

At provision, many devices failed to meet the International Society for Prosthetics and Orthotics standards for safety, workmanship, and client satisfaction. Structured interviews revealed dissatisfaction with function (26%), workmanship (34%), and fit (56%). At follow-up, 45% of clients considered their new device comfortable or somewhat comfortable, while 36% reported discomfort or pain. Among those with a previous prosthesis, 80% preferred it. Usage rates of the provided device were low; 81% reported not very often, rarely, or never, whereas of 280 clients with a previous device, 87% reported often or always using it. At the three-month follow-up, 29% of clients were using a previous device observed at the camp as unused, broken, painful, or poorly fitting, suggesting many individuals rely on an inadequate or dangerous prosthesis.

These findings indicate that previously reported shortcomings in quality and satisfaction of prostheses delivered at intensive fitting camps persist, and raise new questions about inclusiveness of patient selection and effectiveness of funding use. Inadequate training and aftercare may exacerbate these issues, increasing the strain on local physical rehabilitation services or leaving vulnerable clients without support. Future camps should be fully integrated with the existing services and should leave behind adequate materials and components for repairs and replacement. Screening of patients for need is essential, as is engagement of the practitioner who will be expected to continue with the care of the patients.

## Introduction

Functional prostheses can greatly enhance the lives of individuals with lower limb amputation by improving mobility, fostering independence and enabling social participation, benefitting overall quality of life [1]–[4]. However, as with many rehabilitation services, the unmet need for prosthetic and orthotic provision is disproportionately high in low-resource settings (LRS), particularly in regions affected by humanitarian disasters, conflicts, or landmine legacies [5], [6].

To address this need, short-term initiatives such as prosthetic and orthotic ‘missions’ or ‘fitting camps’ aim to provide mobility assistive devices to large numbers of individuals in intensive sessions, often in LRS, on a similar principle to vaccination camps. These camps are distinct from mobile services operated by local prosthetic providers [7], as they are typically funded and delivered by international organisations, such as Non-Governmental Organisations (INGOs).

Although they are well-intentioned, the International Society for Prosthetics and Orthotics (ISPO) has raised concerns about these missions, focusing on the standard of device design, quality of fabrication, and adequacy of aftercare [8]. These concerns highlight the potential harm caused by raising clients’ expectations without sustainable care plans, which can negatively affect both individuals and existing local services. The broader challenges of sustainable prosthetic provision are underscored by ISPO’s definition of *appropriate technology* in P&O. This concept, established in recommendations from 1996 to 2001, emphasises prosthetic rehabilitation services that ensure proper fit and alignment based on sound biomechanical principles, meet individual needs and remain affordable and sustainable within the country. Notable examples include the Polypropylene Technology limbs introduced by the International Committee of the Red Cross (ICRC) [9]–[11] and the Jaipur Foot and Jaipur Limb [12].

Cambodia provides a compelling case study in external support for landmine clearance and physical rehabilitation services following the Vietnam War, Cambodian Civil War, and Khmer Rouge regime. The Cambodian government has historically struggled to manage a multitude of often incoherent external health interventions [13]. However, INGOs operating in the country have successfully established P&O services with a clear strategy of transitioning management to domestic authorities. By 2023, Cambodia’s physical rehabilitation network comprised 11 centres run by the Royal Cambodian Government and INGOs, all using the standardised Polypropylene Technologies devices.

At the time of writing, the Jaipur Foot Organisation reports having held 104 ‘on-the-spot’ fitment camps since 1975 across 42 countries in Asia, Africa, and Central and South America [14]. In March 2023, the Jaipur Limb Organisation organised an intensive prosthetic limb fitting camp in Cambodia, from 14-31/03/2023, near Sisophon, Banteay Meanchey Province, near the Cambodian-Thai border. Following concerns about the quality and functionality of the Jaipur limbs [15], [16], the Cambodian Mine Action and Victim Assistance Authority (CMAA) requested an assessment of the prosthetic devices fitted during the camp. In response to this request, we evaluated both the quality of the delivered devices and the clients’ satisfaction.

## Methods

### Study design

To comprehensively evaluate both device quality and client satisfaction, a multiple-methods approach was employed. This included a standardised assessment of device delivery, completed jointly by a prosthetist and the client immediately after the fitting, followed by an invitation to participate in a second interview three months later. For clients who consented to be followed up, telephone interviews were conducted to gather insights into their experiences and satisfaction with the device in a community setting. We were given approval by the mission organisers to assess the clients after discharge, but not to intervene.

The study design was tailored to the Cambodian context, specifically to assess the outcomes and user experiences at an intensive fitting camp. Ethical approval for data collection was granted by the Cambodian National Ethics Committee for Health Research (NECHR, ref160) and for analysis by the University of Southampton Ethics and Research Governance Office (ERGO, ref102303).

### Stage 1: Device quality assessment tool

A device assessment checklist was completed by ISPO-certified clinicians and clinical educators, developed from the device delivery assessment procedure used at established Cambodian PRCs (SK, TH, CH). It evaluated five factors associated with device quality, using the guiding principle: “If a student prosthetist submitted this, would it be acceptable?” Additionally, the checklist included seven factors related to client acceptance. Finally, the clinicians provided further comments, allowing a summary of their observations and any additional detailed comments made by the clients.

To ensure a standardised and efficient approach to data collection, a bespoke Microsoft Form was developed, allowing data to be collated in a Microsoft Excel spreadsheet for analysis. A team of nine people administered the quality assessment on the day of device fitting in the camp, in pairs which included one senior prosthetist with >15yrs experience, and a second with minimum 5yrs experience.

### Stage 2: Follow-up structured telephone interviews

For consenting clients who could be contacted, additional interviews were conducted by telephone three months after device delivery. These employed a semi-structured approach, with questions associated with status and frequency of use of their new device, the ongoing use of their previous device where applicable, and their preferences for a future device. Interviews were carried out by five of the same ISPO-Certified Prosthetists who carried out Stage 1. Responses were documented directly into a Microsoft Form.

### Data Analysis

Data was analysed by a team of researchers who were independent from data collection (AC, MDH, AD), to ensure objectivity and minimise researcher bias.

#### Stage 1: Device quality assessment tool

Data from the standardised assessment tool was coded into positive and negative responses, and analysed using descriptive statistics, including summed scores across the clinician-assessed questions (quality), client-assessed questions (satisfaction), and the total. Open-ended responses were analysed using content analysis [17]. This is an established method for categorising text and summarising the frequency of response categories numerically. Each response was a treated as a separate unit of analysis and assigned a descriptive code based on their content. These codes were then organised into a coding frame, which was used to analyse related response. New codes were added if existing ones did not adequately capture the content. The final coding frame was used to systematically code the data and determine the frequency of responses. Throughout the analysis, the coding process and tentative categories were discussed between MDH & AD and revised to enhance the credibility of the findings. Consensus was high, with only minor modifications following these discussions.

#### Stage 2: Follow-up structured telephone interviews

Data from the telephone follow-up interviews was coded into categories representing device status, use level and preference, and analysed using descriptive statistics.

## Results

### Stage 1: Device quality assessment

Device delivery assessments were completed for 542 devices received by 532 individual clients, of whom 330 completed a follow-up telephone survey between 25/05 and 12/06/2023 (Table 1). Nine devices were orthoses which were left out of the subsequent analysis, leaving 525 individual clients at delivery and 324 at follow-up, who were prescribed with 533 prosthetic limbs. The median age was 59 years (inter-quartile range IQR 54-64). Clients were predominantly male, with transtibial amputations due to mine or other weapon/ordnance injury. The median time since last device delivery was 5 years (IQR 3-11).

**Table 1:**
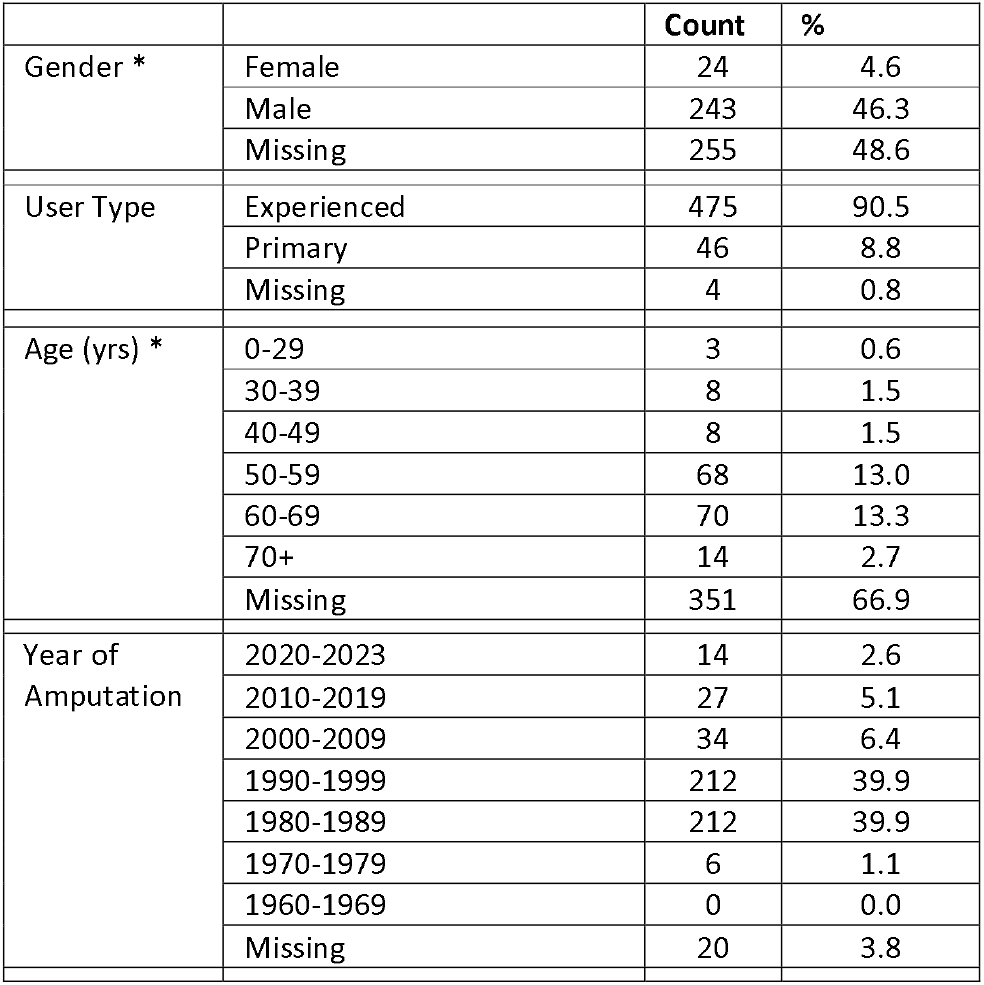
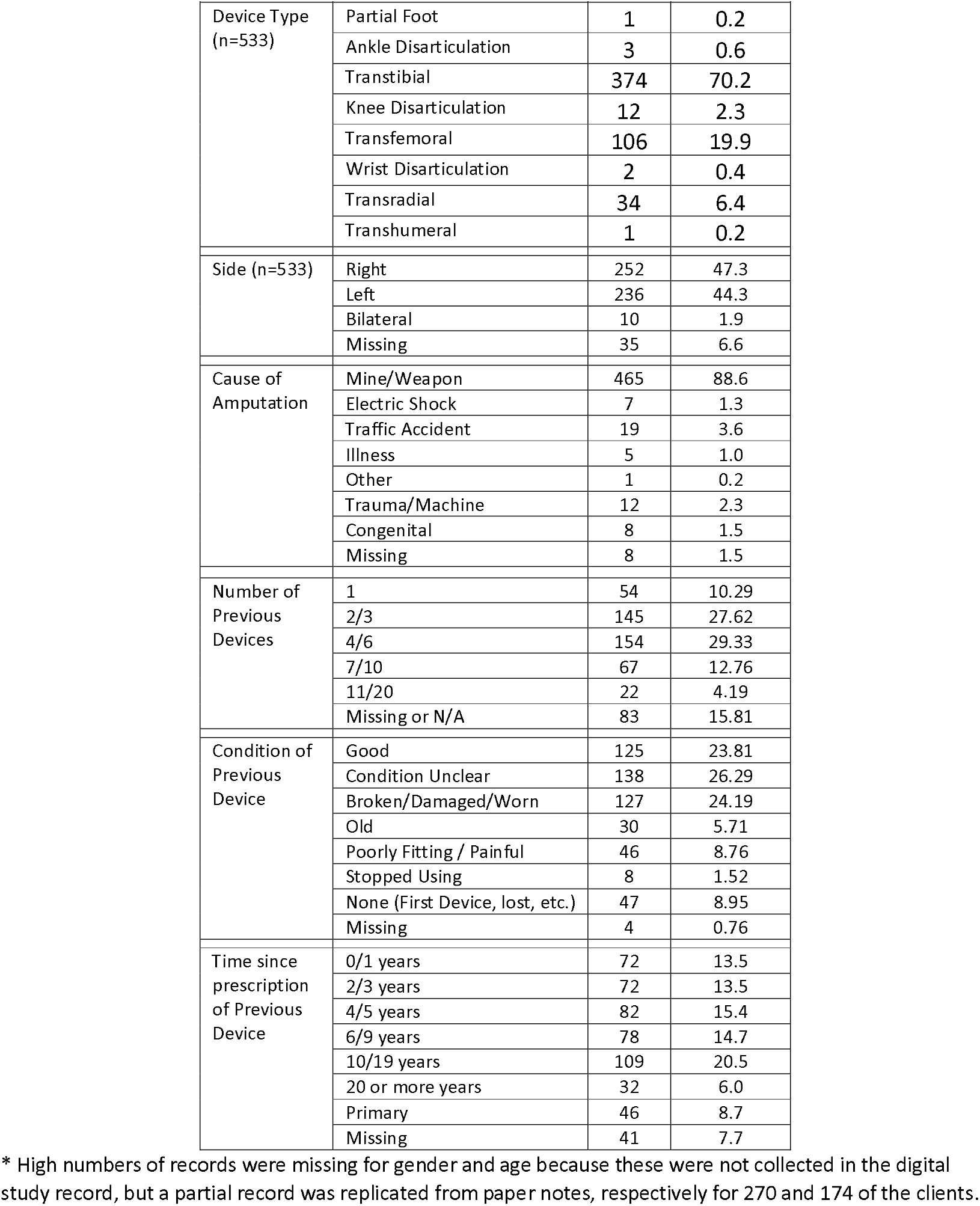
Client demographic and health characteristics, and description of their previous prosthetic device.

Device delivery assessments were completed for all except six participants (Figure 1, Table 2), and identified several areas in which the devices did not meet safety, workmanship and client satisfaction criteria. Examples of poor workmanship and cosmetic appearance are shown in Figure 2.

**Table 2:** Prosthetist assessments and client feedback at the point of device delivery.

| Prosthetist Assessments (n=529): | Yes | No | % Yes | % No |
| --- | --- | --- | --- | --- |
| P1. Is the general workmanship appropriate? | 15 | 507 | 3 | 97 |
| P2. Is the socket fit of device correct? (Weight bearing, total contact, shape, socket fitting, comfortable?) | 45 | 474 | 9 | 91 |
| P3. While standing, is the height of the device correct? | 315 | 203 | 61 | 39 |
| P4. While walking, is the dynamic gait & alignment correct? (pole vertical, foot full contact, stable, any major gait deviations? ..) | 44 | 462 | 9 | 91 |
| P5. After doffing the device, is the residual limb in good condition?* | 407 | 107 | 79 | 21 |
| Client Feedback (n=528): | Yes | No | % Yes | % No |
| C1. While standing, walking and sitting is the device comfortable and stable? | 333 | 188 | 64 | 36 |
| C2. While walking and sitting, is the suspension or straps adequate? | 272 | 248 | 52 | 48 |
| C3. Can the patient use the device independently? (donning, doffing, walking, ...) | 495 | 27 | 95 | 5 |
| C4. Does the device's function meet the patient needs? Correct prescription? | 285 | 229 | 55 | 45 |
| C5. Is the device's cosmetic appearance appropriate? | 47 | 475 | 9 | 91 |
| C6. Is the client informed about device care? | 35 | 487 | 7 | 93 |
| C7. Is the client satisfied with new device and willing to give up their current device? | 185 | 297 | 38 | 62 |

\* This was assessed after approximately ten minutes owing to constraints of the number of clients to see within the study period, which may be insufficient to detect a risk of residual limb tissue injury. This would usually be assessed after approximately one hour of device use (i.e. gait training) in a conventional clinic.

**Figure 1:**
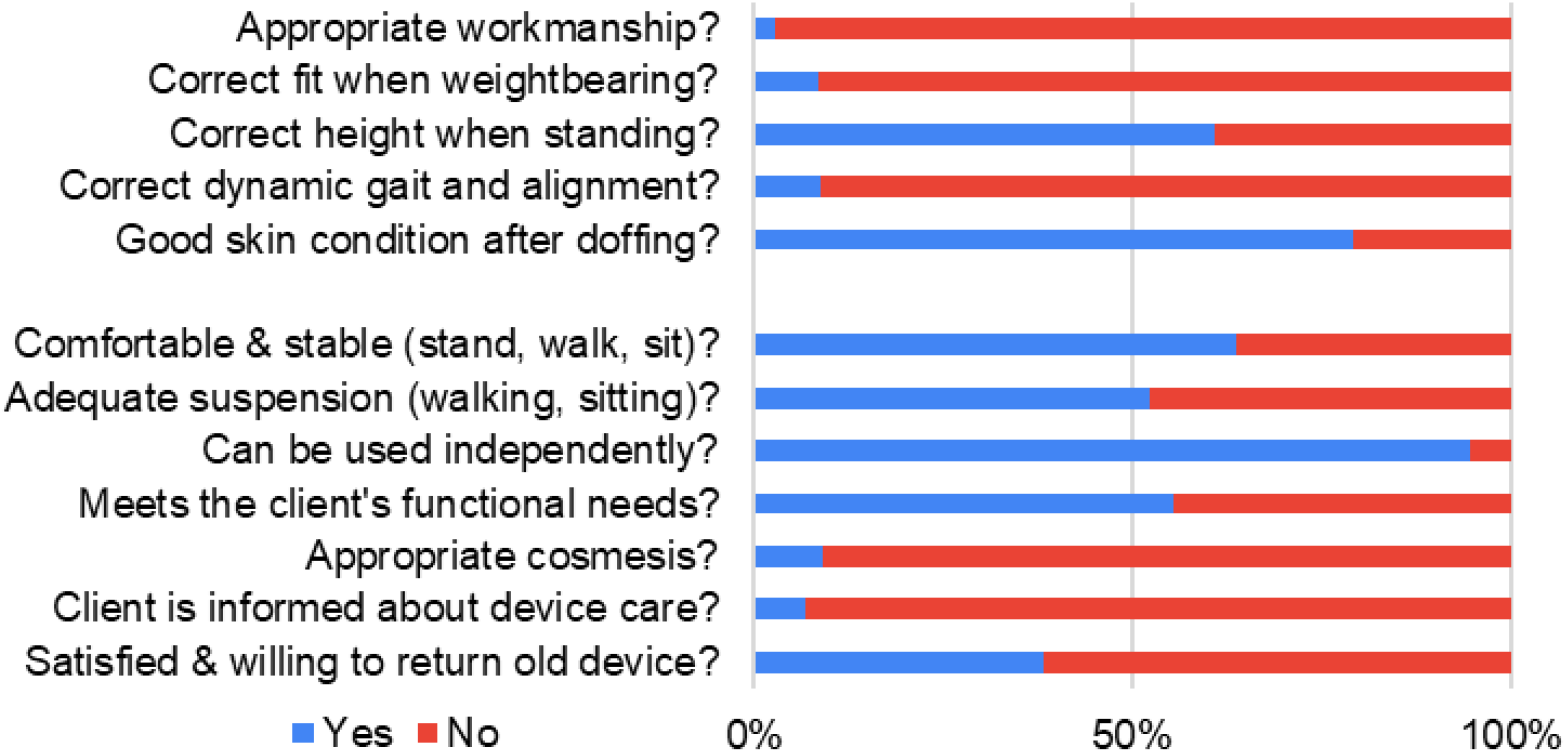
Prosthetist assessments and client feedback at the point of device delivery.

**Figure 2:**
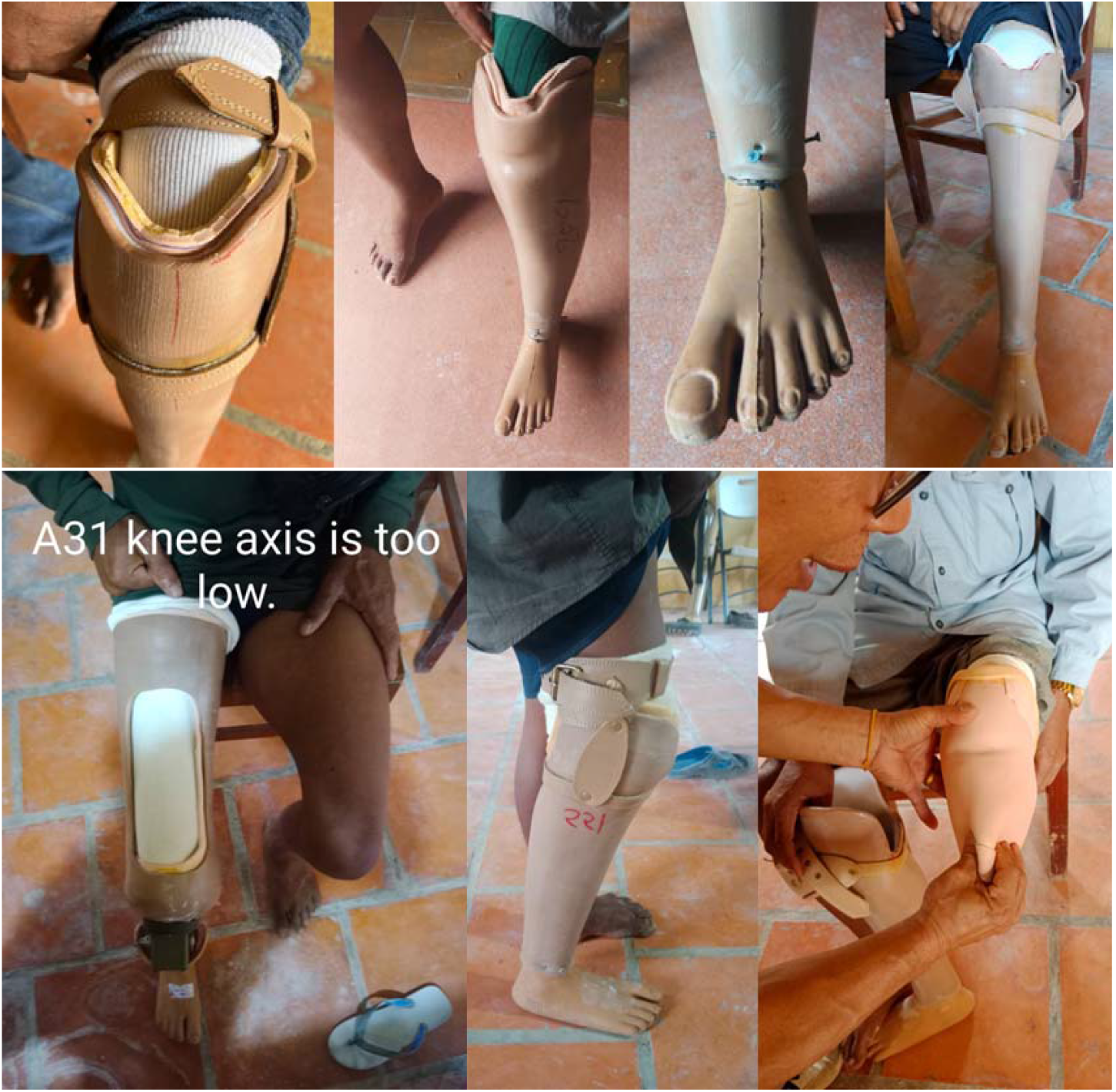
photographs of example transtibial prosthetic devices showing poor device design quality and fabrication workmanship. Devices and liners were observed to have thick walls, with roughly finished brims. Some had exposed screws at the ankle, and poor cosmesis including pen marks, excess adhesive, and visible transition between cosmesis and foot. Others had poor alignment of the componentry such as knee axis height or socket alignment.

Three scores were calculated by summing the positive responses in each i) prosthetist assessment, ii) client assessment and iii) combined prosthetist and client assessment (Figure 3, Appendix Table 5). The mean prosthetist assessment score was 1.58/5, the mean client assessment score was 3.35/7, and the mean overall score was 4.93/12. As such, at Stage 1 the clients’ opinion was more positive than the prosthetists’, though nearly 2/3 of clients were not sufficiently satisfied with the new device to give up their previous device.

**Figure 3:**
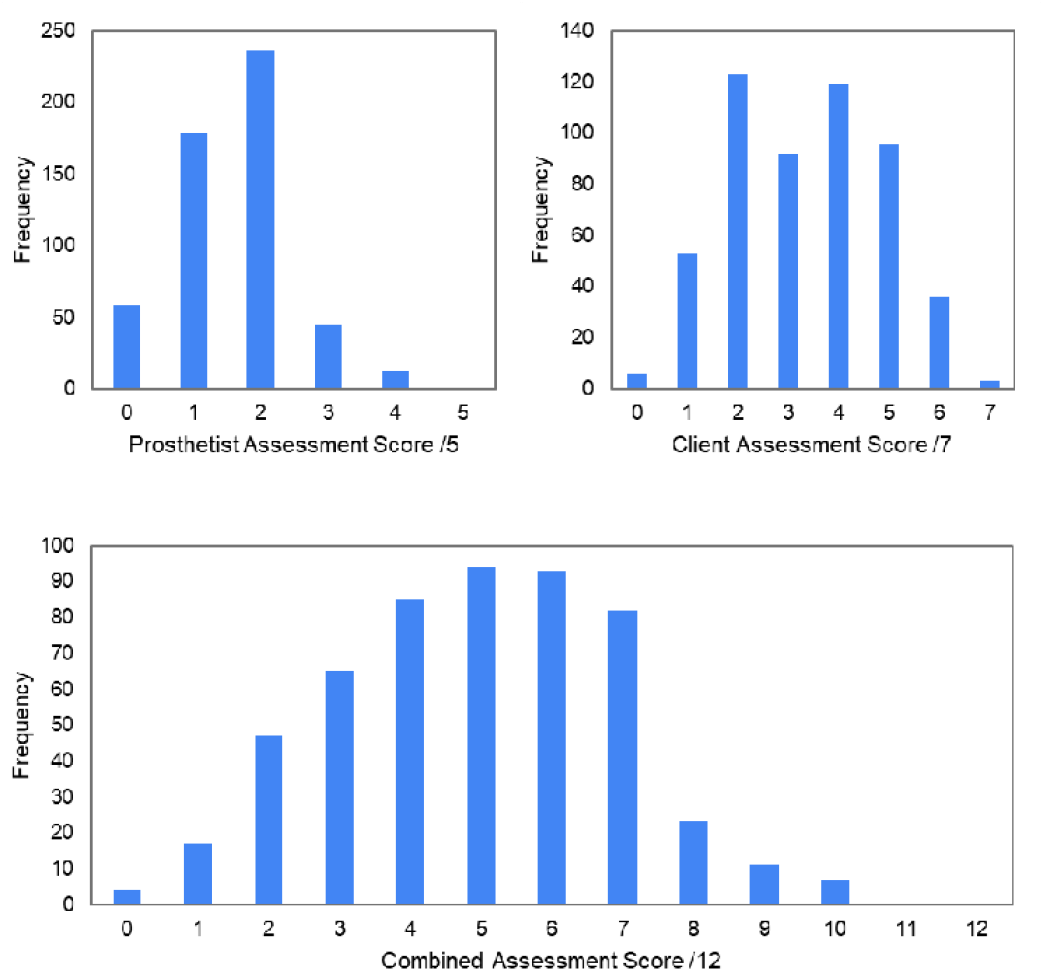
Prosthetist assessments and client feedback at the point of device delivery.

### Content Analysis of Open-Ended Comments

Content analysis of clinicians’ responses to the open ‘Further Comments’ question at device delivery (Table 3) revealed a widespread range of reasons for dissatisfaction and inadequacies associated with use and mobility, cosmesis and finish, device fit and client experience, consistent with the quality assessment and photographic evidence (Figure 2). Clinicians reported gait deviations (n=139, 26%), wrong height (n=110, 21%) and poor alignment (n=66, 12%). The majority also reported observing, or the client reporting, a socket fit that was either too loose (n=230, 43%) or too tight (n=67, 13%), and poor finishing (n=175, 34%) including pen marks, exposed rivets, sharp trim lines and unadhered cosmeses, though some commented upon good experience with the device’s light weight (n=51, 10%).

**Table 3:** Content analysis of structured interview responses at device delivery.

| Category |  | Code | Count |
| --- | --- | --- | --- |
| Use / mobility | Positive | Client can walk, do activities of daily living | 4 |
|  | Negative | Gait deviation | 139 |
|  | Negative | Prosthesis wrong height | 110 |
|  | Negative | Poor alignment | 66 |
|  | Negative | Difficulty donning and doffing | 14 |
|  | Negative | Difficult / unstable walking | 12 |
|  | Negative | Knee functions poorly, or hard to control | 12 |
|  | Negative | Noise when walking | 6 |
|  | Negative | Difficulty wearing device | 5 |
|  | Negative | Requires training/practice | 5 |
|  | Negative | Cannot walk unaided | 3 |
|  | Negative | Walks with knee locked | 2 |
|  | Negative | Cannot walk at all | 1 |
| Cosmesis / Finish | Positive | Client likes appearance | 2 |
|  | Negative | Poor cosmesis / finish in general | 175 |
|  | Negative | Trim line sharp or uneven | 74 |
|  | Negative | Cuff suspension visible through long pants | 5 |
|  | Negative | Rivet/cuff not secure | 2 |
|  | Negative | Socket attachment | 2 |
|  | Negative | Visible when sitting | 2 |
|  | Negative | Rivet showing | 1 |
|  | Negative | Rubber not stuck | 1 |
|  | Negative | Prosthesis not strong enough | 1 |
| Socket Fit | Positive | Good socket fit in general | 19 |
|  | Negative | Too loose | 230 |
|  | Negative | Poor socket fit in general | 143 |
|  | Negative | Too tight/pressure | 67 |
|  | Negative | Incorrect trim line height | 29 |
|  | Negative | Inadequate suspension | 10 |
| Experience | Positive | Light weight | 51 |
|  | Negative | Socket causes pain | 19 |
|  | Negative | Socket not smooth | 10 |
|  | Negative | Prosthesis uncomfortable | 6 |
|  | Negative | Prosthesis too heavy or bulky | 2 |
|  | Negative | Straps too tight | 1 |

### Stage 2: Follow-Up Interviews

Analysis of Stage 2 interview questions at 3-months follow-up showed that 138 (45%) of clients considered their new device as comfortable or somewhat comfortable, compared to 112 (36%) not comfortable or painful (Table 4, Appendix Figure 5 top). Five reported the new device was broken.

**Table 4:** Categoric responses to questions at three month follow-up, regarding device condition, use level and preference.

|  |  | Count | % |
| --- | --- | --- | --- |
| Condition of new device | Comfortable | 108 | 33.3 |
|  | Somewhat comfortable | 34 | 10.5 |
|  | Not comfortable | 115 | 35.5 |
|  | Broken | 6 | 1.9 |
|  | Does not fit / function | 3 | 0.9 |
|  | Has poor cosmesis | 1 | 0.3 |
|  | Painful | 2 | 0.6 |
|  | Easy to use | 1 | 0.3 |
|  | Do not use | 54 | 16.7 |
| Which device can you walk further and faster on?* | New device | 39 | 13.0 |
|  | Previous device | 241 | 80.1 |
|  | Neither | 21 | 7.0 |
| Which device do you prefer?* | New device | 41 | 13.9 |
|  | Previous device | 237 | 80.3 |
|  | Neither | 17 | 5.8 |
| Use level of the new device | Always | 61 | 18.8 |
|  | Often | 0 | 0 |
|  | Short Periods / Some Days | 127 | 39.2 |
|  | Rarely | 97 | 29.9 |
|  | Never / Cannot use / Broken | 34 | 10.5 |
|  | Does not have | 0 | 0 |
|  | Time not stated / Other | 5 | 1.5 |
| Use level of the previous device | Always | 185 | 57.1 |
|  | Often | 63 | 19.4 |
|  | Short Periods / Some Days | 1 | 0.3 |
|  | Rarely | 15 | 4.6 |
|  | Never / Cannot use / Broken | 20 | 6.2 |
|  | Does not have | 35 | 10.8 |
|  | Time not stated / Other | 4 | 1.2 |
|  | Missing | 1 | 0.3 |

\* only includes those who could compare; excludes those who did not have a previous device, e.g. primary patients.

**Table 5:**
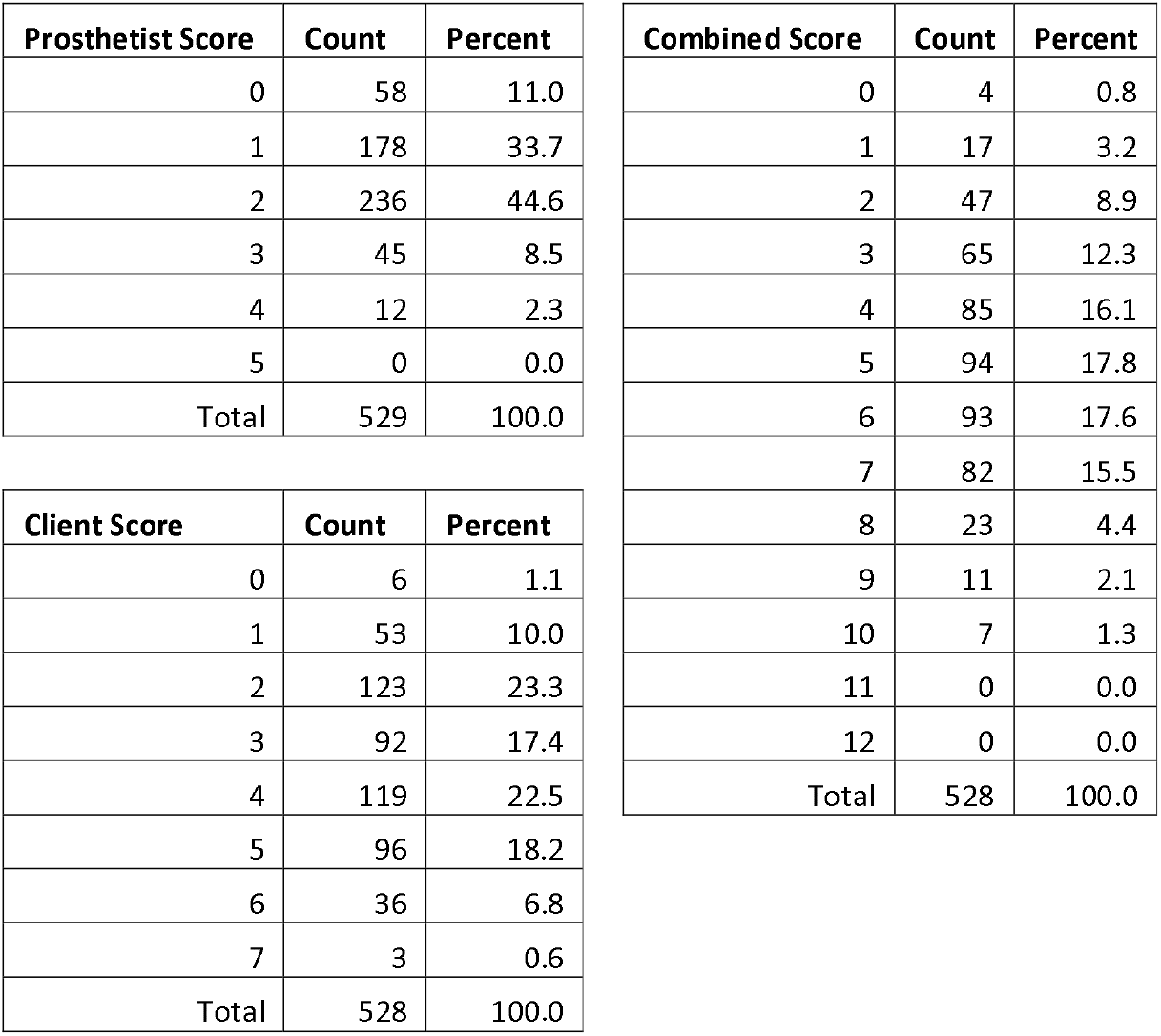
Prosthetist, client and combined assessment scores.

Of the clients who could compare between their new and previous devices (i.e. excluding primary patients) 39 said they could walk further and faster with the new device compared to 233 who selected their previous device, and 21 reported both were the same (Table 4, Appendix Figure 5 bottom). Considering preference, 233 (80%) selected their previous device.

Considering usage, 258 respondents (81%) reported not very often, rarely, almost never or never using their new device, and 61 (19%) reported using their new device always. Asked about their previous device, of 280 clients who still had a previous device, 244 (87%) reported often or always using it.

Relatively little difference was observed between categories of the clients’ previous device condition at the camp visit (Stage 1) and their usage of both the new and previous devices at 3-months follow-up (Stage 2). A high proportion of clients (80-92%) who had a previous device had reverted to using it most or all of the time across all previous device condition categories (Figure 4 bottom). Of clients whose previous device was damaged or worn out, only 28% were using their new device all the time, and of the clients who had no previous device, 87% either used their new device for short periods, rarely or never (Figure 4 top).

**Figure 4:**
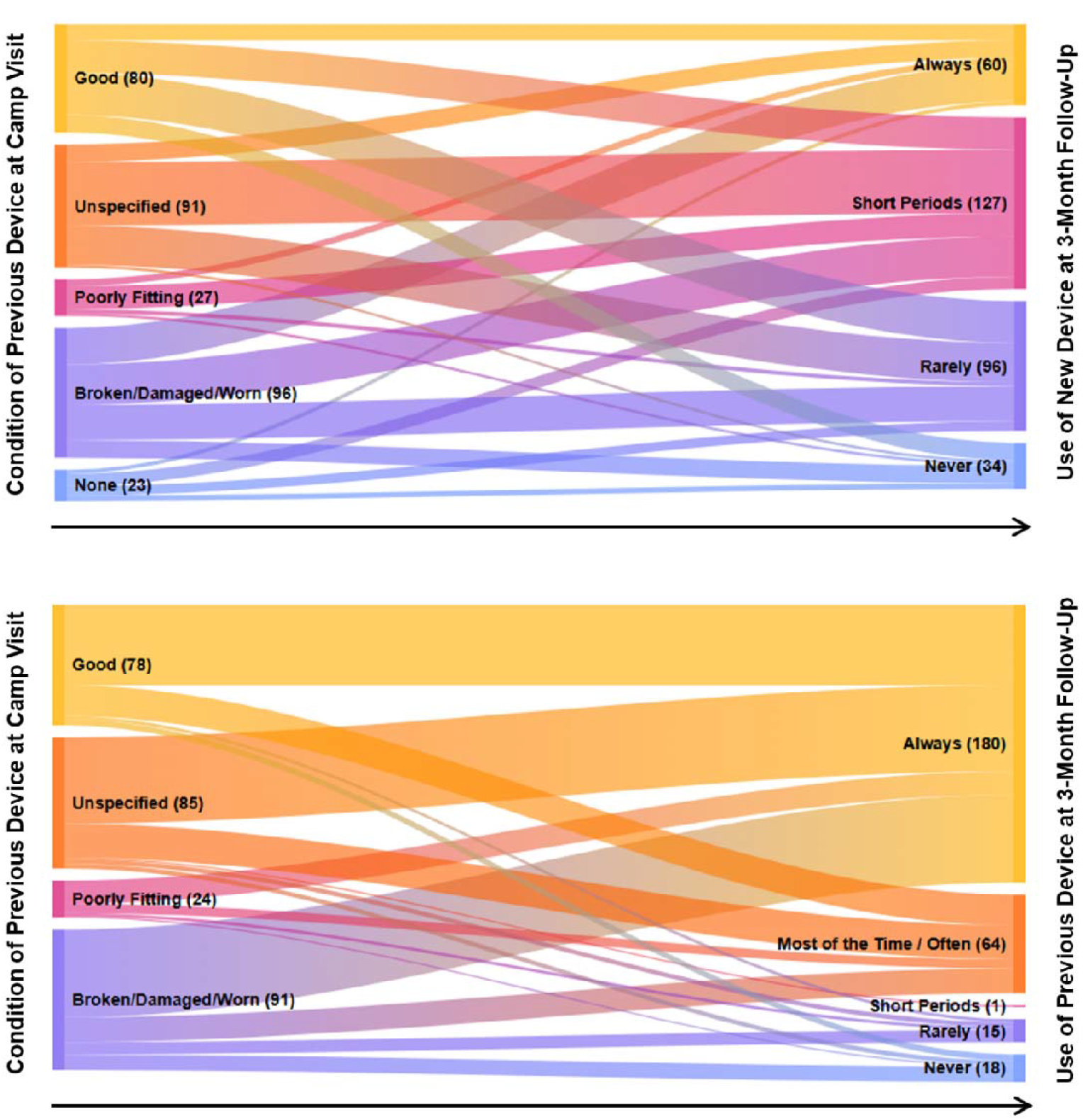
Mapping from the clients’ reported previous device status to their level of use of their new device (top), and their previous device (bottom) at 3 months follow-up. Bracketed numbers are counts.

**Figure 5:**
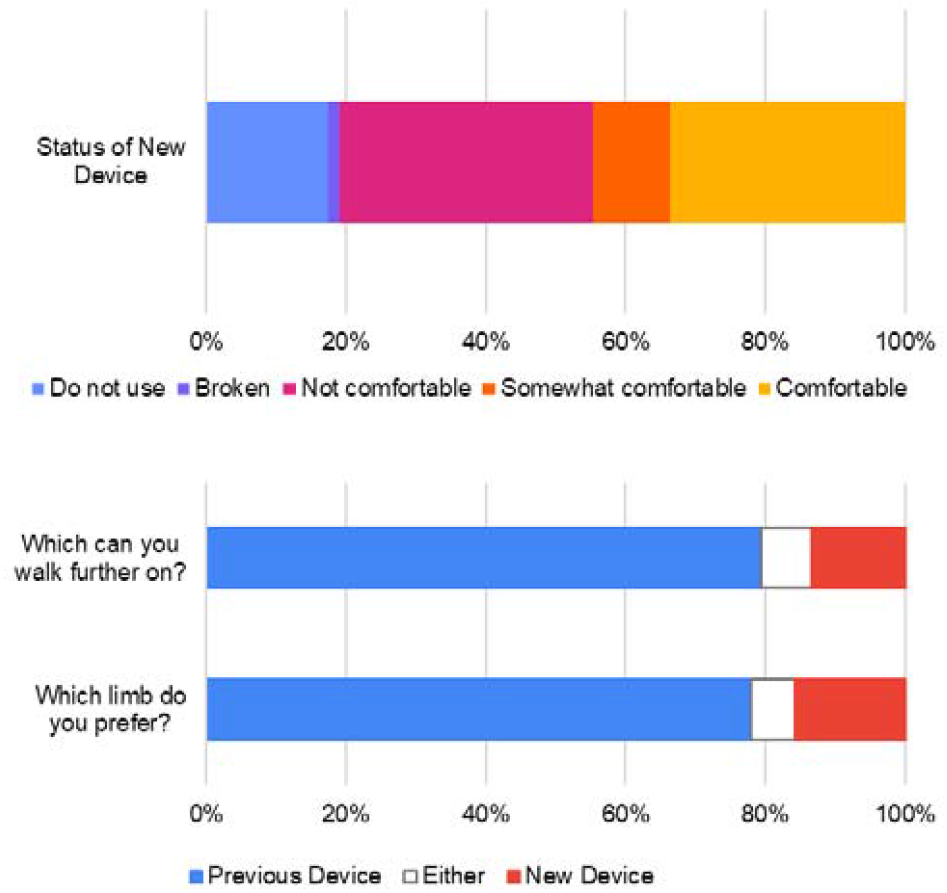
Client report of device status (top), and device preference (bottom) at 3 months follow-up. For latter two categories, percentages calculated using clients who reported having both devices, unbroken.

**Figure 6:**
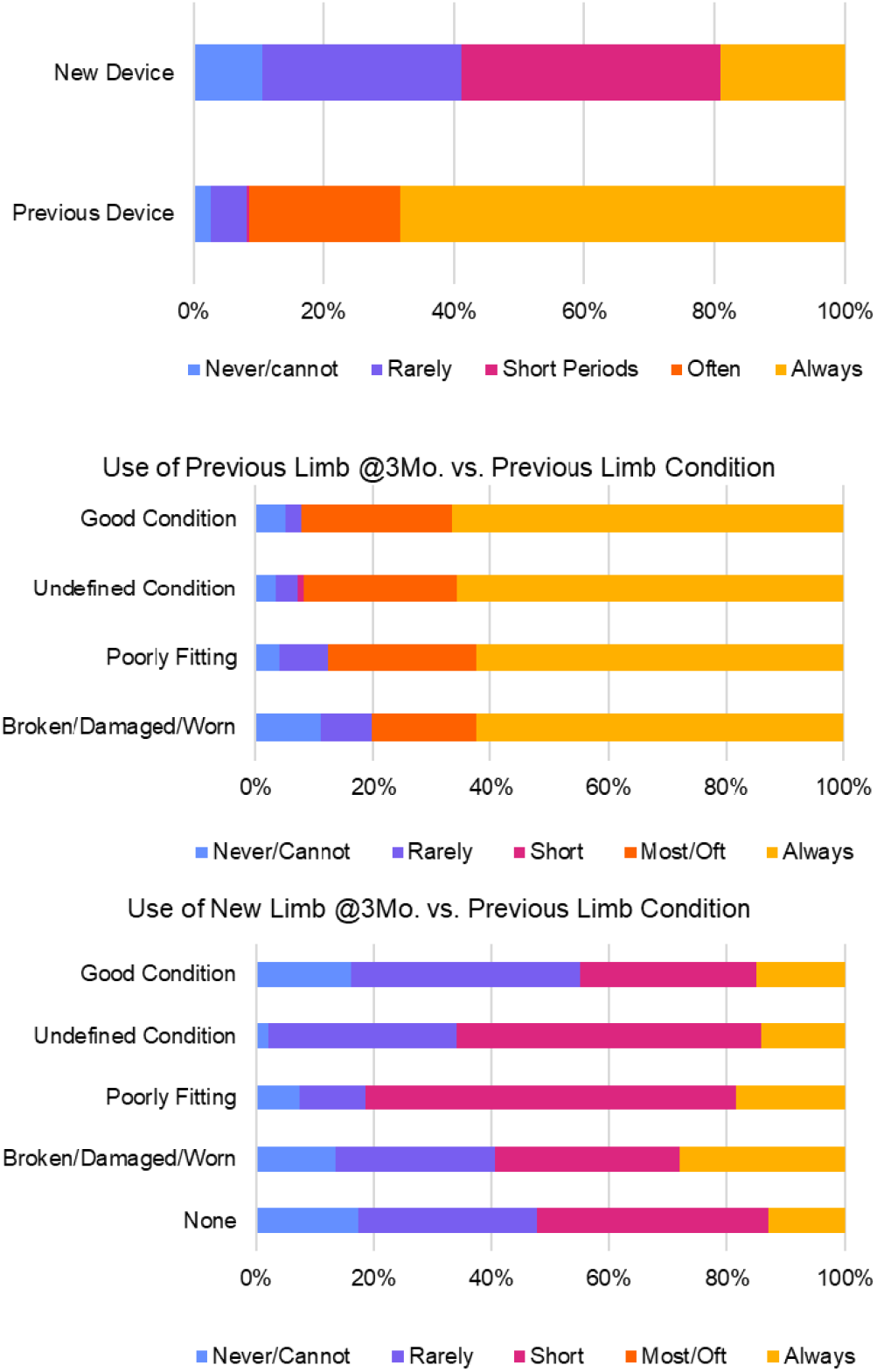
Client report of previous and new device usage at 3 months follow-up, for whole group completing follow-up (top), and broken down by groups according to previous limb condition (middle and bottom).

## Discussion

### Key findings and comparison to prior studies

The present study was conducted on the request of the Royal Government of Cambodia’s Mine Action and Victim Assistance Authority (CMAA) following the organisation of a camp providing Jaipur limbs. The outcomes did not meet the benchmarks set by the International Society for Prosthetics and Orthotics (ISPO), which include targets for 95% compliance, 90% satisfaction, 60% good socket fit (including maximum 10% needing socket change), 90% acceptable alignment and 90% good craftsmanship [18], [19]. In conventional Cambodian PRCs, devices failing any of the twelve quality or satisfaction checks at delivery would be re-worked, especially considering those criteria which might lead to poor function and potential injury. Additionally, the outcomes were lower than those reported in multiple low-resource settings (Cambodia, Vietnam, El Salvador and Tanzania) in ISPO/USAID-supported initiatives [20].

While the Jaipur Foot meets many mechanical, social, and cultural-specific needs, it has faced criticism for its weight, lack of manufacturing standardisation and limited durability, particularly for individuals with higher body weights [21], [22]. The present study’s findings suggest that quality and satisfaction remain key concerns for these devices, aligning with reports from more than 20 years ago. In 2004, ISPO-affiliated authors conducted a ∼3-year follow-up of Jaipur devices in three countries (Honduras, Uganda, and India) at transtibial [15] and transfemoral [16] levels. They identified poor craftsmanship in 56% of transtibial and 86% of transfemoral devices, primarily due to poor fit, alignment, socket wall adequacy, and leg length discrepancy. Although satisfaction and compliance were reported to be good for transtibial device users (85% and 94%, respectively) and moderate in transfemoral device users (58% and 65%, respectively) fewer than half the participants could walk more than 1 km and many reported discomfort and pain. These 2004 papers had a considerably longer follow-up than the present study, however the observed quality shortcomings associated with device fit, length and alignment cannot reasonably be expected to improve without further intervention. Indeed, those shortcomings that might be managed-such as accommodating socket looseness by wearing socks-may be undesirable in the warm climate experienced for most of the year in Cambodia and many other LRS [15].

Furthermore, a significant proportion of clients opted to retain their old devices, and a large proportion had reverted to using their previous device within three months of receiving the new prosthesis. This suggests that, despite their initially more positive assessment, the clients’ experience over time aligned with the prosthetists’ assessments at fitting. It is possible that Cambodia’s political history instilled a culture of “*hierarchy and ranking, deference and command*” [23], which may be more prominent in rural communities [24]. As a result, individuals may be unaccustomed or reluctant to voice concerns about their physical rehabilitation care and prosthetic devices, especially when these are provided free of charge. This is reflected in the present study, where clients reported some level of satisfaction with their new devices while also noting discomfort, imperfect fit, and concerns about cosmesis and workmanship.

More broadly, these findings raise questions about patient selection for the camp, and specifically whether all attendees had a genuine need for a new device or if their previous prosthesis remained adequate. An important characteristic of the present study’s population is that those with a previous device reported having used it for a median of 5 years (Table 1). Our previous analysis of clients of a regular NGO-run physical rehabilitation service in Cambodia showed a median time to device repair of 727 days, or two years [11], and this may indicate that a notable proportion of the clients had less good access to physical rehabilitation service. However, at least 125 clients were identified to have a previous device described as in good condition at the point of new device delivery, representing 23.8% of the full cohort. 72 clients were using devices that had been delivered in the last year, of whom 17 received their devices prior to the camp during 2023, making them less than three months old.

In contrast, 211 clients (40.2% of the cohort) were identified to have a previous device that was reported to be unused, broken, painful and/or poorly fitting (too loose or tight). However, 94 clients in this subset, representing 29.0% of those completing Stage 2, were identified to be using their previous device after 3 months, suggesting a significant number of people could be living with an inadequate or dangerous prosthesis. Of the 138 clients who had a previous device whose condition could not be ascertained, 90 completed Stage two and of them the majority (78 clients, 87%) had returned to using the previous device. As such, both the percentages of clients who had a well-functioning alternative device prior to the camp (indicating poor client selection), and those who had returned to using an inadequate device 3 months after it (indicating poor device quality), are likely to be under-estimates.

### Limitations

The study used a bespoke assessment tool, which considered quality and satisfaction against criteria previously defined by ISPO but was not standardised or previously validated. This led to some heterogeneity in completion of the questions, and some potentially valuable client criteria were not captured, notably gender. Furthermore, potential biases arise due to the circumstances of the assessment and the need to collect large amounts of data in a timely manner, and to assess all clients. The data was collected by a relatively large number of certified prosthetist assessors potentially leading to some subjectivity, and the study did not include a process of recording or checking. A significant proportion of the study participants (38%) could not be contacted for the Round 2 telephone interviews and so were considered lost to follow-up. However, no substantial difference was observed in the Stage 1 device assessment at delivery between these clients and the full group.

### Summary and outlook

Central to Dr Sethi’s vision for the Jaipur Limb was the demystification of prosthetic knowledge and the simplification of technology, enabling the establishment of camps that not only provide devices but also train local artisans to fabricate, adjust, and repair them [12]. Beside the large number of camps and delivered devices reported by the Organisation [14], it has been stated that “*Somewhere down the line, the number of amputees fitted at these camps overtook the concept of imparting training to the local artisans, and it all boiled down to a game of numbers*” [25]. This has been attributed in part to replacement of the initially successful beaten aluminium socket fabrication technique with use of thermoplastic from HDPE pipes, which was quicker and cheaper but heavier and more difficult to achieve the desired alignment and fit. Further, the ISPO-affiliated 2004 follow-up studies of these devices reported that the *“material and components are of high technical standard and could provide a low-cost possibility, but improvement is needed. The utilisation of manpower is unacceptable. The untrained, so-called technicians are unable to adapt a prosthesis to an amputation stump with a functional result even with more sophisticated materials and components. A recognised prosthetics training is required to ensure proper use of materials and correct alignment of the prosthesis*.” [16]. In initial discussions, the investigators offered to review, provide training needs analysis and training, and the offer was not taken up.

The present study identifies that the previously reported concerns regarding the quality and satisfaction with devices delivered at intensive limb fitment camps may persist. The study also presents new evidence which calls into question their patient selection, and whether this represents an inclusive model of care or an effective use of funding, despite the best of intentions to provide care to those who may not be able to access it. Previous research has identified that device durability and access to repairs and servicing are reported as issues of top priority to people in LRS who use prostheses and orthoses [4], [26]. Inadequacies in training and follow-up care may have widespread consequences of burdening local physical rehabilitation services if they exist, or leave vulnerable clients without support. Future camps should be fully integrated with the existing services and should leave behind adequate materials and components for repairs and replacement. Screening of patients for need is essential, as is engagement of the practitioner who will be expected to continue with the care of the patients.

## Data Availability

All data produced in the present study are available upon reasonable request to the authors

## Appendices

## References

[1] D. Wyss, S. Lindsay, W. L. Cleghorn, and J. Andrysek, “Priorities in lower limb prosthetic service delivery based on an international survey of prosthetists in low- and high-income countries,” Prosthet. Orthot. Int., vol. 39, no. 2, pp. 102–111, 2015, doi: 10.1177/0309364613513824.

[2] B. F. Mundell, H. M. Kremers, S. Visscher, K. M. Hoppe, and K. R. Kaufman, “Predictors of Receiving a Prosthesis for Adults With Above-Knee Amputations in a Well-Defined Population,” PM&R, vol. 8, no. 8, pp. 730–737, 2016, doi: 10.1016/j.pmrj.2015.11.012.

[3] R. Stuckey, P. Draganovic, M. M. Ullah, E. Fossey, and M. P. Dillon, “Barriers and facilitators to work participation for persons with lower limb amputations in Bangladesh following prosthetic rehabilitation,” Prosthet. Orthot. Int., vol. 44, no. 5, pp. 279–289, 2020, doi: 10.1177/0309364620934322.

[4] N. Ramstrand, A. Maddock, M. Johansson, and L. Felixon, “The lived experience of people who require prostheses or orthoses in the Kingdom of Cambodia: A qualitative study,” Disabil. Health J., vol. 14, no. 3, p. 101071, 2021, doi: 10.1016/j.dhjo.2021.101071.

[5] J. Borg, A. Lindström, and S. Larsson, “Assistive technology in developing countries: A review from the perspective of the Convention on the Rights of Persons with Disabilities,” Prosthet. Orthot. Int., vol. 35, no. 1, pp. 20–29, 2011, doi: 10.1177/0309364610389351.

[6] A. J. Ikeda, A. M. Grabowski, A. Lindsley, E. Sadeghi-Demneh, and K. D. Reisinger, “A scoping literature review of the provision of orthoses and prostheses in resourcelimited environments 2000-2010. Part one: Considerations for success,” Prosthet. Orthot. Int., vol. 38, no. 4, pp. 269–286, 2014, doi: 10.1177/0309364613500690.

[7] M. Farrar, Y. R. Niraula, and W. Pryor, “Improving access to prosthetic services in Western Nepal: a local stakeholder perspective,” Disabil. Rehabil., vol. 45, no. 7, pp. 1229–1238, 2023, doi: 10.1080/09638288.2022.2057599.

[8] C. Harte, “Prosthetic orthotic missions: Ethics and efficacy,” Prosthet. Orthot. Int., vol. 46, no. 5, p. 407, 2022, doi: 10.1097/PXR.0000000000000186.

[9] J. S. Jensen and S. Heim, “Evaluation of polypropylene prostheses designed by the International Committee of the Red Cross for trans-tibial amputees,” Prosthet. Orthot. Int., vol. 24, no. 1, pp. 47–54, 2000, doi: 10.1080/03093640008726521.

[10] S. Weerasinghe, A. Aranceta-Garza, and L. Murray, “Efficacy of rehabilitation after provision of ICRC lower limb prostheses in low-income and middle-income countries: A quantitative assessment from Myanmar,” Prosthetics Orthot. Int., no. November, 2023, doi: 10.1097/pxr.0000000000000300.

[11] A. S. Dickinson et al., “Understanding repair and replacement of prosthetic limbs using routinely-collected data: a retrospective study over three decades in Cambodia,” medRxiv, pp. 1–27, 2024, doi: 10.1101/2024.10.15.24315396.

[12] P. K. Sethi, “Technological choices in prosthetics and orthotics for developing countries,” Prosthet. Orthot. Int., vol. 13, no. 3, pp. 117–124, 1989, doi: 10.3109/03093648909079418.

[13] F. Bourdier, “Health inequalities, public sector involvement and malaria control in Cambodia,” Sojourn J. Soc. Issues Southeast Asia, vol. 31, no. 1, pp. 81–115, 2016, doi: 10.1355/sj31-1c.

[14] “BMVSS Jaipur Foot.” [Online]. Available: www.jaipurfoot.org. [Accessed: 16-Dec-2024].

[15] J. S. Jensen, J. G. Craig, L. B. Mtalo, and C. M. Zelaya, “Clinical field follow-up of high density polyethylene (HDPE)-Jaipur prosthetic technology for trans-tibial amputees,” Prosthet. Orthot. Int., vol. 28, no. 3, pp. 230–244, 2004, doi: 10.3109/03093640409167755.

[16] J. S. Jensen, J. G. Craig, L. B. Mtalo, and C. M. Zelaya, “Clinical field follow-up of high density polyethylene (HDPE)-Jaipur prosthetic technology for trans-femoral amputees,” Prosthet. Orthot. Int., vol. 28, no. 2, pp. 152–166, 2004, doi: 10.1080/03093640408726700.

[17] P. Mayring, Qualitative Content Analysis: A Step-by-Step Guide. London: SAGE Publications, 2021.

[18] J. S. Jensen, R. Nilsen, and J. Zeffer, “Quality benchmark for trans-tibial prostheses in low-income countries,” Prosthet. Orthot. Int., vol. 29, no. 1, pp. 53–58, 2005, doi: 10.1080/17461550500085147.

[19] J. S. Jensen, W. Raab, J. Fisk, C. Hartz, A. Saldana, and C. Harte, “Quality of polypropylene sockets for trans-tibial prostheses in low-income countries,” Prosthet. Orthot. Int., vol. 30, no. 1, pp. 45–59, 2006, doi: 10.1080/03093640600568336.

[20] J. Steen Jensen and S. Sexton, “Appropriate Prosthetic and Orthotic Technologies in Low Income Countries (2000-2010),” USAID/ISPO, Brussels, 2010.

[21] A. P. Arya and L. Klenerman, “The Jaipur foot,” J. Bone Jt. Surg. - Ser. B, vol. 90, no. 11, pp. 1414–1416, 2008, doi: 10.1302/0301-620X.90B11.21131.

[22] I. Huber et al., “Epidemiological study of failures of the Jaipur Foot,” Disabil. Rehabil. Assist. Technol., vol. 13, no. 8, pp. 740–744, 2018, doi: 10.1080/17483107.2017.1369593.

[23] D. Chandler, Facing the Cambodian Past: Selected Essays 1971-1994. Chiang Mai: Silkworm Books, 1998.

[24] K. Un, “State, society and democratic consolidation: The case of Cambodia,” Pac. Aff., vol. 79, no. 2, pp. 225–245, 2006, doi: 10.5509/2006792225.

[25] R. Bhargava, “The Jaipur foot and the ‘Jaipur Prosthesis,’” Indian J. Orthop., vol. 53, no. 1, pp. 5–7, 2019, doi: 10.4103/ortho.IJOrtho_162_18.

[26] L. Magnusson and G. Ahlström, “Patients’ Satisfaction with Lower-limb Prosthetic and Orthotic Devices and Service delivery in Sierra Leone and Malawi,” BMC Health Serv. Res., vol. 17, no. 1, pp. 1–13, 2017, doi: 10.1186/s12913-017-2044-3.

